# How much unplanned hospital activity is for people in their last year of life and how did the Covid-19 pandemic impact this? A 10-year population study for England (2014-2023)

**DOI:** 10.64898/2026.10.23.26366298

**Authors:** Joanna M Davies, Julia Verne, Thomas Bolton, Jadene Lewis, Rachel L Chambers, Anna E Bone, Katherine E Sleeman, the CVD-COVID-UK/COVID-IMPACT Consortium

## Abstract

**Objectives:** The aim of this study is to investigate the impact of the Covid-19 pandemic on the proportion of unplanned hospital use that is for people in their last year of life in England, and to examine changes over time from 2014 to 2023.

**Design:** A whole population study using secondary analysis of linked routinely collected health data.

**Setting:** England between 1^st^ January 2014 and 31^st^ December 2023.

**Participants:** Adults aged 18+ years using unplanned hospital care.

**Main outcome measures:** Unplanned hospital activity for people in their last year of life.

**Results:** Between 2014-2023, Emergency Department (ED) visits for people in their last year of life increased from 900,295 (6.2% of total) to 1,089,365 (6.1% of total); ambulance conveyances to the ED increased from 696,395 (17.3% of total) to 769,545 (19.5% of total); unplanned hospital admissions increased from 835,960 (18.0% of total) to 889,920 (16.4% of total); and unplanned hospital bed days increased from 11,180,815 (29.2% of total) to 11,491,915 (28.2% of total).

The Covid-19 pandemic was associated with an increase in the proportion of unplanned hospital admissions that were for people in their last year of life, with the largest increase observed among people living in more deprived areas (after adjusting for age and sex).

**Conclusions:** These findings show that people in their last year of life account for a substantial proportion of demand for acute hospital and ambulance services. The changes observed during the Covid-19 pandemic underscore the need to consider end-of-life care needs when planning and delivering acute hospital services during periods of exceptional pressure. Improving the quality and coordination of end-of-life care across hospital and community care services should be a key consideration in efforts to meet the needs of an ageing population while managing pressures on the health system.

**What is already known on the topic?:**

- For people in their last year of life, unplanned hospital use is common and increases in the months before death.
- Very little is known about how the proportion of unplanned hospital use for people who are in their last year of life has changed over time and in response to the Covid-19 pandemic.

**What this study adds:**

- Approximately one-third of unplanned hospital bed days are for people in their last year of life indicating that care of the dying is a core activity for hospitals.
- The increase in the proportion of unplanned hospital admissions that were for people in their last year of life during the Covid-19 pandemic, highlights the need to consider end-of-life care needs when planning services during periods of exceptional pressure.

## Background

Internationally, hospitals face growing operational strain, with emergency admissions reflecting broader pressures within the health and social care system.^1–3^ Towards the end of life, unplanned hospitalisations are common and increase in the months before death.^4–7^ For people living with terminal illness, emergency hospital visits towards the end of life can be distressing; some are appropriate but many could be avoided through better coordinated care and community support.^8–10^

In 2014, Clark et al published a study using Scottish data, showing that one-third of patients in hospital at any one time were likely to die within a year.^11^ This important study highlighted that care of dying patients is ‘core business’ for hospitals and added to calls for palliative and end of life care to be given greater priority in hospital settings. Public health monitoring data for England for 2021 showed that 15% of unplanned hospital admissions and 31% of unplanned bed days were for people in their last year of life.^12^ The Covid-19 pandemic was associated with both increased mortality and major disruption to end-of-life care, including changes in place of death.^13,14^ However, we know very little about how the proportion of unplanned hospital activity that is for people in their last year of life has changed in response to the pandemic.

Most people want to avoid hospitalisation in their last months of life. Yet, in the UK 63% of people have at least one Emergency Department (ED) visit in their last 3 months of life^5^ and of the 12 billion pounds spent each year on healthcare for people in their last year of life, 81% is spent on hospital care.^15^ A better understanding of the extent to which unplanned hospital use is accounted for by people in their last year of life, and how this has changed over time, would help to focus attention on the need for better care for dying people in the community and identify ways to alleviate pressure on hospitals.

The aim of this study is to investigate the impact of the Covid-19 pandemic on the proportion of unplanned hospital use that is for people in their last year of life in England, and to examine changes over time from 2014 to 2023.

## Methods

### Study design

This is a whole population study using secondary analysis of linked data assets for England.^16^ We accessed individual-level anonymised, national, linked, electronic health records for England in NHS England’s Secure Data Environment service for England, made available via the British Heart Foundation (BHF) Data Science Centre’s CVD-COVID-UK/COVID-IMPACT Consortium. Data processing and linkage are described elsewhere.^16^

### Data sources

We used seven linked datasets, including:

1. Hospital Episode Statistics (HES) Accident and Emergency (A&E)
2. Hospital Episode Statistics (HES) Emergency Care Dataset (ECDS) –used to supplement HES A&E information on ‘mode of arrival’
3. Hospital Episode Statistics (HES) Admitted Patient Care (APC)
4. Mortality data
5. BHF Health Data Science (HDS) Curated Asset for Demographics^17^ - for consolidating commonly used patient information (e.g., sex, date of birth, Lower Super Output Area of residence, etc.) from multiple data sources (General Practice Extraction Service (GPES) Data for Pandemic Planning and Research, HES APC/Out Patients/A&E, Deaths, Vaccine status, etc.), to maximise completeness and quality.
6. HES APC Continuous Inpatient Spell (CIPS) dataset (a HDS curated asset)^18^ – for linking in valid discharge dates where these were missing from the HES APC data

### Study population

Adults aged 18+ years in England using unplanned hospital care between 1st Jan 2014 and 31st December 2023.

### Variables and analysis

The analysis is at activity-level (not patient-level), and includes the following types of unplanned hospital activity:

i. Emergency Department (ED) visits
ii. Ambulance conveyance to ED
iii. Unplanned hospital admissions
iv. Unplanned hospital bed days

For ED visits and ambulance conveyances, we included all ED visits, which may include a small number of cases that originate from within the hospital. For unplanned hospital admissions and bed days, we restricted the analysis to those classified as an ‘emergency admission’, therefore excluding ‘elective admissions’.^19^

For each type of unplanned hospital activity, we calculate the number and proportion of overall activity that was for people in their last year of life, for each quarter of each year between 2014 and 2023 and depict this in a line graph. For ED visits, we describe the activity separately for visits in hours and visits out-of-hours (evenings (after 6pm and before 8am), weekends and bank holidays).

For unplanned hospital admissions, to understand changes before, during, and after the Covid-19 pandemic, we describe the proportion of activity that is for people in their last year of life in 3 time periods:

- During the Covid-19 pandemic (including wave 1 from March 2020 to May 2020, and wave 2 from September 2020 to April 2021^20^): 1st March 2020 – 31st April 2021
- Pre-pandemic comparison period: 1st March 2018 – 31st April 2019
- Post-pandemic comparison period: 1st March 2022 – 31st April 2023

We describe how the proportion of unplanned hospital admissions that are for people in their last year of life varies across these time periods by age at visit, sex, and area-based deprivation. Area-based deprivation is used as a proxy for individual level socioeconomic position based on the neighbourhood (lower super output area (LSOA)) of residence at the time of the hospital admission using the Index of Multiple Deprivation 2019.^21^

To better understand the association between area-based deprivation and the probability of being in the last year of life, and how this association varied across the three study periods, we fitted a modified Poisson regression model including deprivation quintile, time period, age, sex, and an interaction between deprivation and time period. We did not adjust for additional health-related characteristics, such as comorbidity, because our aim was to describe age- and sex-adjusted socioeconomic inequalities rather than estimate the direct effect of deprivation independent of confounders or mediators through which deprivation may operate.

We used a modified Poisson approach with robust standard errors to estimate adjusted relative risks, as these are generally more readily interpretable than odds ratios from logistic regression. Analyses were restricted to complete cases, with individuals excluded if data were missing for any variable included in the model.

Given the exceptionally large study population, the purpose of the modelling was descriptive rather than inferential. We did not seek to test the statistical significance of the interaction term, as even very small differences would be expected to achieve conventional levels of statistical significance in a dataset of this size. Instead, the model was used to adjust for differences in age and sex composition across deprivation groups and time periods, allowing meaningful comparison of adjusted outcome probabilities. We therefore present marginal predicted proportions derived from the interaction model. These were estimated with age fixed at the median age of the study population and averaged across the observed distribution of sex, providing age- and sex-adjusted estimates of the probability of being in the last year of life for each deprivation quintile within each study period.

We also describe the proportion of unplanned hospital admissions that are for people in their last year of life for each Integrated Care Board (ICB). The ICB is based on the patients’ place of residence at the time of the admission and we use the ICB boundaries from 2022.^22^

The analysis using data in NHS England’s Secure Data Environment (SDE) for England was performed according to a pre specified analysis plan published on GitHub, along with the analysis code https://github.com/BHFDSC/CCU110).

This study is reported in accordance with the STROBE guideline and the REporting of studies Conducted using Observational Routinely-collected health Data (RECORD) statement.^23^

### Patient and Public Involvement

As part of the data application process, we consulted with two patient and public involvement (PPI) partners who advised on the aims, objectives and overall plan for the analysis, and helped us to plan the substantive PPI element of the project. Following this advice, we held a two-hour workshop with ten PPI members to discuss the initial findings from the study. The workshop informed the interpretation of the key messages and the dissemination plan.

## Results

Between 2014-2023, Emergency Department (ED) visits increased from 900,295 (6.2% of total) to 1,089,365 (6.1% of total); ambulance conveyances to the ED increased from 696,395 (17.3% of total) to 769,545 (19.5% of total); unplanned hospital admissions that were for people in their last year of life increased from 835,960 (18.0% of total) to 889,920 (16.4% of total); and unplanned hospital bed days increased from 11,180,815 (29.2% of total) to 11,491,915 (28.2% of total) (figure 1 and 2 and supplementary file 1, sheets 1-4).

**Figure 1:**
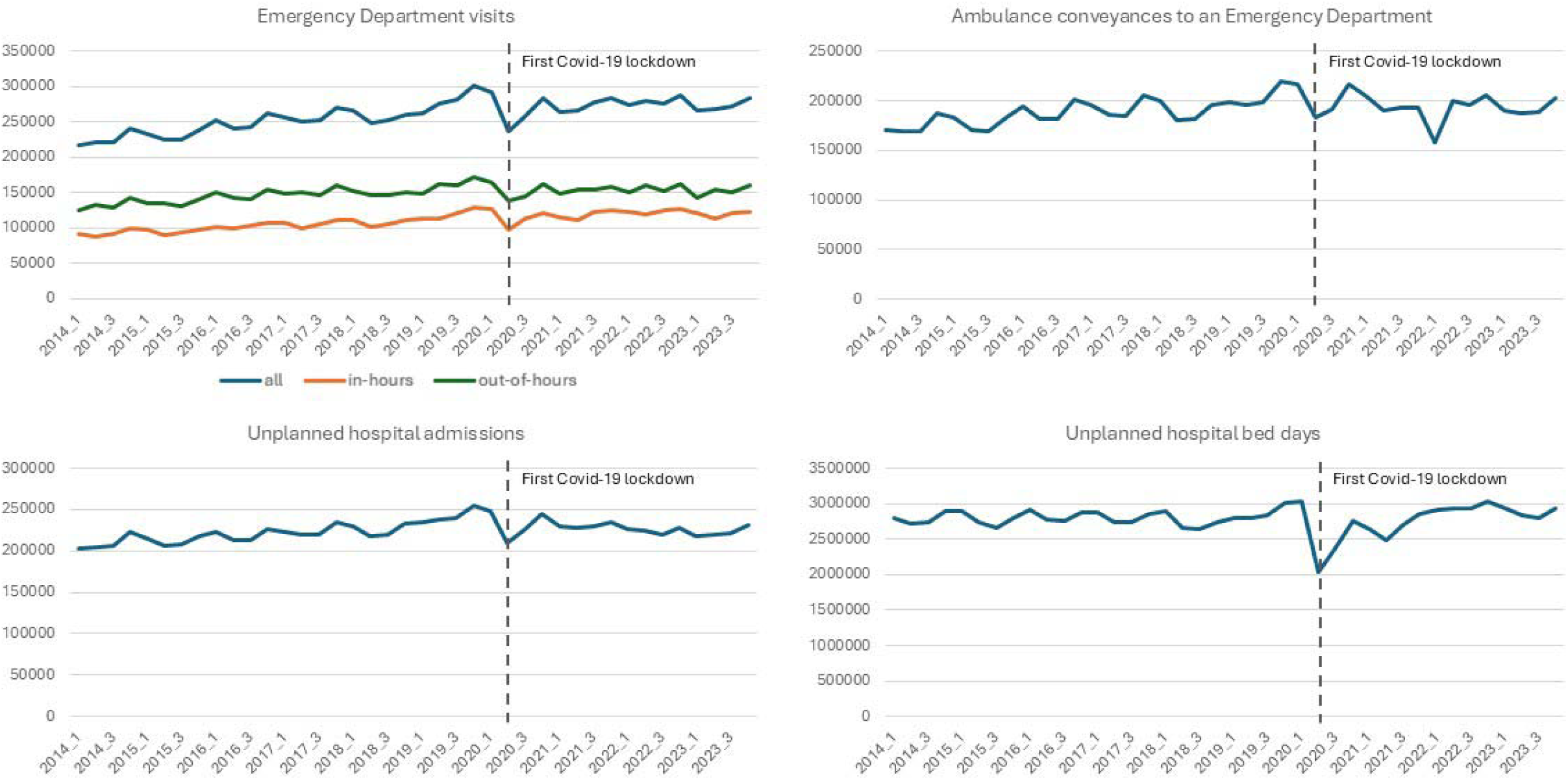
Quarterly count of unplanned hospital activity that was for people in their last year of life in England between 2014 and 2023.

**Figure 2:**
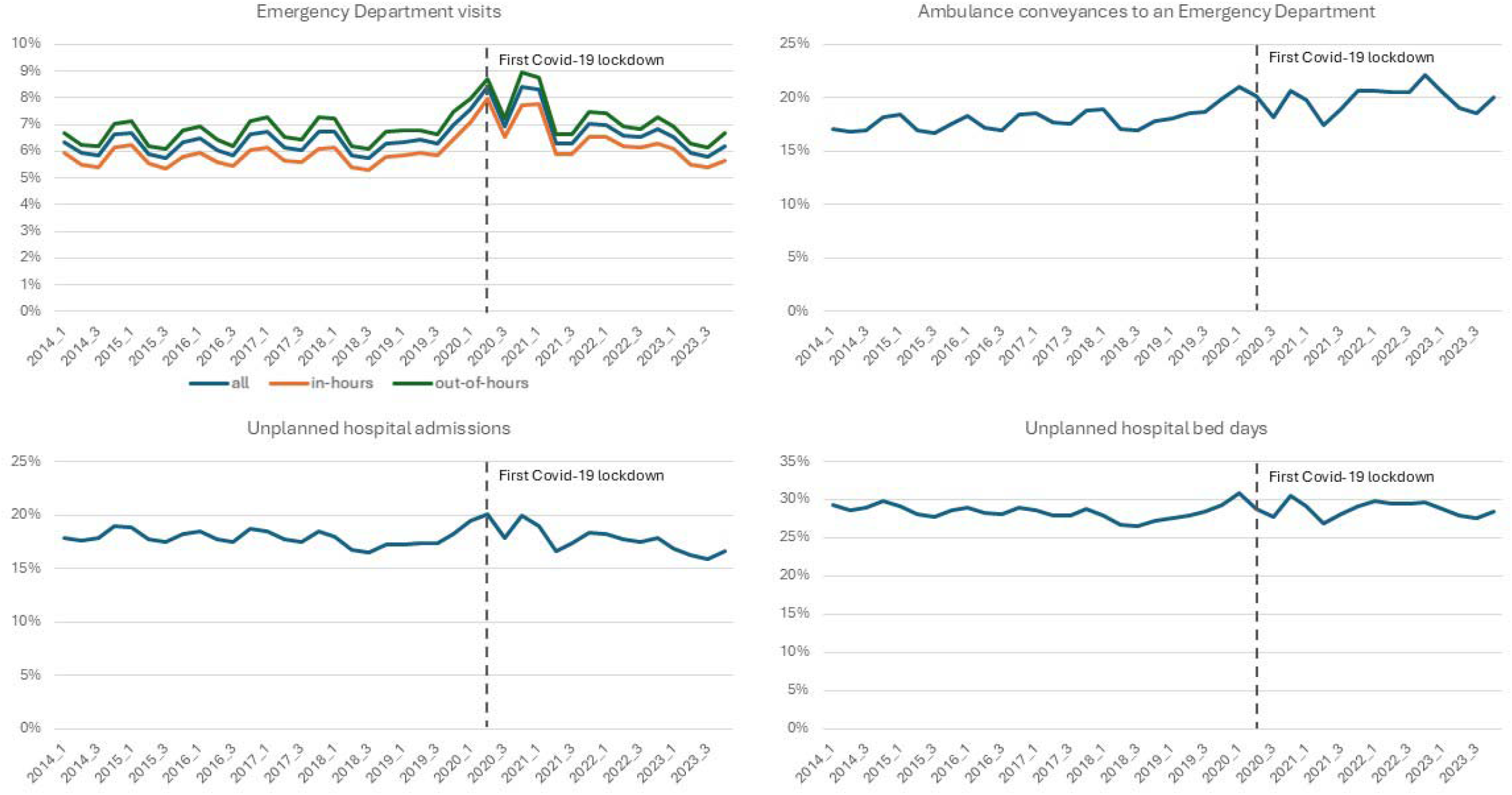
Quarterly proportion of unplanned hospital activity that was for people in their last year of life in England between 2014 and 2023.

For all types of unplanned hospital activity, we see a reduction in the count of activity corresponding with the first Covid-19 lock down in quarter 2 of 2020.

We see a consistent seasonal pattern across the different types of unplanned activity, with small increases in the number and proportion of activity that is for people in their last year of life seen in quarter 4 (Oct-Dec) and quarter 1 (Jan-Mar), corresponding with the higher mortality expected during the winter period (supplementary file 1, sheet1-4).

### Unplanned hospital admissions before, during and after the Covid-19 pandemic

During the Covid-19 pandemic, a higher proportion of unplanned hospital admissions were for people in their last year of life across all groups (apart from those with missing data) and this reduced in the post-Covid-19 pandemic period (table 1). The proportion of unplanned hospital admissions that were for people in their last year of life increased with age and was higher for men consistently across the three time periods.

**Table 1:** Number and proportion of unplanned hospital admissions for people in their last year of life (lyol), by sex, age, and deprivation, and across time periods (pre, during and post the Covid-19 pandemic*)

|  | Pre Covid-19 |  |  | During Covid-19 |  |  | Post Covid-19 |  |  |
| --- | --- | --- | --- | --- | --- | --- | --- | --- | --- |
|  | total | lyol | % in lyol | total | lyol | % in lyol | total | lyol | % in lyol |
| Median age | 64 | 80 |  | 65 | 80 |  | 65 | 80 |  |
| (IQ range) | (44 to 79) | (70 to 87) | - | (45 to 79) | (70 to 87) | - | (44 to 79) | (70 to 87) | - |
| **Age** |  |  |  |  |  |  |  |  |  |
| <60 | 2727425 | 115565 | 4.2% | 2402245 | 116970 | 4.9% | 2553530 | 111280 | 4.4% |
| 60-69 | 842205 | 134705 | 16.0% | 768535 | 135625 | 17.6% | 850470 | 134110 | 15.8% |
| 70-79 | 1125895 | 259185 | 23.0% | 1021680 | 265810 | 26.0% | 1121435 | 264635 | 23.6% |
| 80+ | 1552760 | 552835 | 35.6% | 1382525 | 543970 | 39.3% | 1426445 | 530200 | 37.2% |
| **Sex** |  |  |  |  |  |  |  |  |  |
| men | 2902375 | 549190 | 18.9% | 2623240 | 562140 | 21.4% | 2762540 | 543855 | 19.7% |
| women | 3344830 | 512485 | 15.3% | 2951290 | 500220 | 16.9% | 3188900 | 496355 | 15.6% |
| other | 990 | 615 | 62.1% | 315 | 15 | 4.8% | 390 | 20 | 5.1% |
| missing | 85 |  | 0.0% | 135 |  | 0.0% | 55 |  | 0.0% |
| **Area-based deprivation\*\*** |  |  |  |  |  |  |  |  |  |
| Quintile 1 | 1499930 | 235710 | 15.7% | 1333510 | 233300 | 17.5% | 1412960 | 223580 | 15.8% |
| Quintile 2 | 1305310 | 217625 | 16.7% | 1168020 | 218690 | 18.7% | 1225780 | 208680 | 17.0% |
| Quintile 3 | 1223775 | 213590 | 17.5% | 1098305 | 213685 | 19.5% | 1170230 | 211780 | 18.1% |
| Quintile 4 | 1145650 | 206955 | 18.1% | 1026390 | 206015 | 20.1% | 1112010 | 206230 | 18.5% |
| Quintile 5 | 1019365 | 185355 | 18.2% | 919375 | 187780 | 20.4% | 993385 | 186885 | 18.8% |
| Missing | 54250 | 3055 | 5.6% | 29380 | 2905 | 9.9% | 37530 | 3075 | 8.2% |

\*During pandemic period (1st March 2020 – 31st April 2021); pre-pandemic period (1st March 2018 – 31st April 2019); post-pandemic period (1st March 2022 – 31st April 2023).
\*\*Quintile 1 is most deprived.

In the descriptive analyses (Table 1), across all three time-periods, the proportion of unplanned hospital admissions attributable to people in their last year of life was highest in the least deprived areas (quintile 5) and the proportion decreased with increasing deprivation.

##### Box 1: A note on interpretation of differences between groups

This analysis is focused on activity, not on individuals. It examines the proportion of all unplanned hospital activity that is for people in their last year of life.

The findings tell us how much of the unplanned workload of hospitals, Emergency Departments and ambulance services is for people in their last year of life. The findings do not indicate how much hospital care individual people or groups receive in their last year of life.

For example, if 37% of admissions among people aged 80 years and over are for people in their last year of life, this means that more than one-third of admissions in this age group are attributable to people who are in their last year of life. It does not necessarily mean that older people receive more hospital care at the end of life than younger people.

The main purpose of this analysis is to describe how much hospital activity is for people in their last year of life and help us to understand what this means for planning hospital, ambulance and palliative care services. The findings are not useful for identifying which population groups have the greatest use of hospital care in the last year of life or have the greatest unmet palliative and end of life care needs.

### Interaction between time period and area-based deprivation

During the Covid-19 pandemic, the predicted proportion of unplanned hospital admissions that were for people in their last year of life increased for all deprivation quintiles, compared to pre- and post-pandemic periods (table 2 and figure 3). The marginal predicted proportions show that after adjusting for age and sex, the increase in the proportion of unplanned hospital admissions that were for people in their last year of life was larger for people living in more deprived areas (table 2).

**Figure 3:**
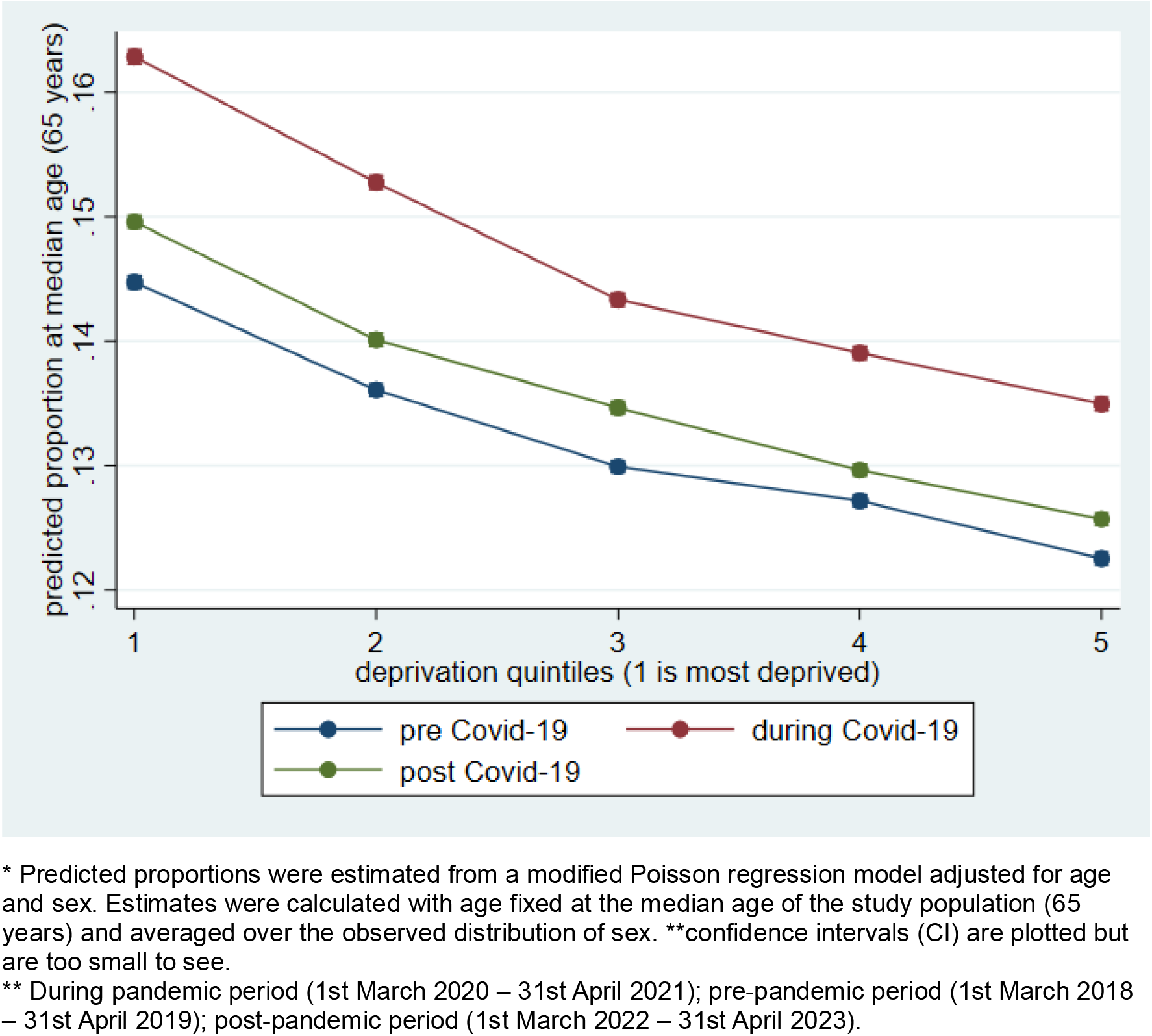
Adjusted predicted proportions* and 95% CI** of unplanned hospital admissions that were for people in their last year of life, by deprivation quintile and time period** (n= 17,653,890)

**Table 2.** Adjusted predicted proportions* of unplanned hospital admissions that were for people in their last year of life, by deprivation quintile and time period** (n= 17,653,890)

|  | pre Covid-19 | during Covid-19 | post Covid-19 |
| --- | --- | --- | --- |
| Quintile 1 (most deprived) | 14.5% | 16.3% | 15.0% |
|  | [14.4, 14.5] | [16.2, 16.3] | [14.9, 15.0] |
| Quintile 2 | 13.6% | 15.3% | 14.0% |
|  | [13.6, 13.7] | [15.2, 15.3] | [14.0, 14.1] |
| Quintile 3 | 13.0% | 14.3% | 13.5% |
|  | [12.9, 13.0] | [14.3, 14.4] | [13.4, 13.5] |
| Quintile 4 | 12.7% | 13.9% | 13.0% |
|  | [12.7, 12.8] | [13.9, 14.0] | [12.9, 13.0] |
| Quintile 5 | 12.3% | 13.5% | 12.6% |
|  | [12.2, 12.3] | [13.4, 13.5] | [12.5, 12.6] |

\* Predicted proportions were estimated from a modified Poisson regression model adjusted for age and sex.
Estimates were calculated with age fixed at the median age of the study population (65 years) and averaged over the observed distribution of sex.
\*\* During pandemic period (1st March 2020 – 31st April 2021); pre-pandemic period (1st March 2018 – 31st April 2019); post-pandemic period (1st March 2022 – 31st April 2023).

The proportion of unplanned hospital admissions that were for people in their last year of life decreased in the post-Covid-19 pandemic period for all groups but remained higher than during the pre-Covid-19 pandemic period.

Compared to the crude rates (table 1), after adjusting for age and sex, we find that the deprivation gradient is reversed (table 2 and figure 3); i.e., with each increase in deprivation (from quintile 5 (least deprived) to quintile 1 (most deprived)) there is an increase in the predicted proportion of unplanned hospital admissions that were for people in their last year of life. Table 2 and figure 3 present the marginal adjusted predicted proportions at median age (65 years), showing consistent step-wise deprivation gradient across the three time periods.

### Integrated Care Boards

Supplementary file 1 (sheet 5) presents the number and proportion of unplanned hospital admissions that were for people in their last year of life for each Integrated Care Board, in each of the three time periods.

## Discussion

Using linked hospital and mortality data, this research examined how much unplanned hospital activity is for adults in their last year of life, in England between 2014 and 2023. We found that people in their last year of life account for a substantial proportion of unplanned hospital care, including 7% of Emergency Department (ED) visits, 19% of ambulance conveyances to ED, 18% of unplanned hospital admissions, and 29% of unplanned hospital bed days. While these proportions were largely stable over time, the absolute numbers increased across all measures. The Covid-19 pandemic was associated with an increase in the proportion of unplanned hospital admissions that were for people in their last year of life, with the largest increase observed among people living in more deprived areas.

Our findings are consistent with previous evidence demonstrating that a substantial proportion of hospital activity is attributable to people in their last year of life. Clark et al. (2014) reported that approximately one in three hospital inpatients in Scottish hospitals were in their last year of life, highlighting the importance of end-of-life care in hospital settings.^11^ However, their study was based on a single inpatient cohort from a single day in Scotland in 2010 and did not examine unplanned hospital use specifically. Similarly, the UK Office for Health Improvement and Disparities reported that 15% of unplanned hospital admissions and 31% of unplanned bed days in England in 2021 (including both adults and children) were among people in their last year of life. Our data extend these findings, providing the first trends over time, including assessment of the impact of the Covid-19 pandemic; in addition, we quantify activity not previously described including ED attendances and ambulance conveyances, demonstrating that people in their last year of life account for a substantial share of urgent and emergency care across the whole pathway.

We found that during the Covid-19 pandemic, the proportion of unplanned admissions that were for people in their last year of life increased, and this increase was largest for people living in the most deprived areas. Our findings align with wider evidence that the Covid-19 pandemic disrupted established patterns of end-of-life care, with increased mortality, changes in care delivery, and unprecedented pressure on both acute and community services.^14,24^ For example, previous research has highlighted substantial changes in where people died during the pandemic, with increases in deaths at home observed across the UK; however, these increases were smallest in the most deprived areas.^24^ Our findings may suggest that, among more deprived populations, a smaller shift towards dying at home was accompanied by a greater contribution of people in their last year of life to unplanned hospital admissions.

In our analysis of the association between deprivation and the probability of being in the last year of life, we found that the deprivation gradient observed in the crude analyses was reversed after adjustment for age and sex. In the unadjusted analyses, a smaller proportion of admissions in the most deprived groups were attributable to people in their last year of life. However, this pattern largely reflects differences in population age structure by deprivation. People living in more deprived areas have a lower life expectancy, whereas populations in less deprived areas tend to be older and therefore contain a larger proportion of people nearing the end of life. As a result, crude comparisons mask the underlying relationship between deprivation and end-of-life status. After accounting for differences in age and sex, we observed a consistent socioeconomic gradient across all study periods, with a higher proportion of unplanned hospital admissions attributable to people in their last year of life among people living in more deprived areas. This finding is consistent with broader evidence of socioeconomic inequalities in health, multimorbidity, and premature mortality across England.^25^

## Strengths and weaknesses of the study

A strength of this study is the use of national, linked hospital and mortality data covering the whole adult population of England over a 10-year period, allowing comprehensive assessment of trends in unplanned hospital activity among people in their last year of life.

The study benefited from high-quality national datasets and highly complete linkage across data sources.^16^ As with all administrative data collected for healthcare delivery rather than research purposes, coding errors, missing data and changes in recording practices may be present; however, these are unlikely to have materially affected the overall findings.

Because the denominator is all unplanned hospital activity, differences between population groups reflect both the amount of activity generated by people in their last year of life and the amount generated by those not in their last year of life. Consequently, a higher proportion of activity attributable to people in their last year of life should not be interpreted as evidence of greater end-of-life care needs or higher hospital utilisation among people who subsequently died. Studies that take all decedents as the denominator are better suited to understanding variation in end-of-life healthcare use and potential unmet need.^5,26^ In contrast, the present analysis is designed to quantify the extent to which care for people in their last year of life contributes to demand on acute hospital services.

Our analyses of the association between deprivation and the probability of being in the last year of life were adjusted for age and sex but not for comorbidities or other potentially important confounders. Comorbidity or other health-related factors may partly mediate the relationship between deprivation and being in the last year of life; therefore, additional adjustment may have attenuated the observed deprivation gradient. However, our aim was to describe socioeconomic differences rather than estimate the direct effect of deprivation independent of other potential causal factors.

## Implications for clinicians and policymakers

The findings have important implications for policy and practice. Approximately one-third of unplanned bed days are attributable to people in their last year of life, indicating that care of people approaching the end of life is a core activity for hospitals and this should be recognised in service planning and workforce development. This sits in stark contrast to the most recent National Audit of Care at the End of Life (NACEL) findings which report that in 2025, just 64% of hospitals in England had a face-to-face specialist palliative care service (doctor and/or nurse) available 8 hours a day, 7 days a week.^27^ Not all hospital inpatients who are in their last year of life will need input from specialist palliative care but the current gaps in provision across the country are indicative of a need for greater investment in these vital services.

Almost one in five ambulance conveyances to the ED involve people in their last year of life, highlighting the important contribution that people living with terminal illness made to the overall caseload of paramedics. Elsewhere research has highlighted the challenges faced by paramedics in providing care to patients with terminal illness often during a crisis.^28^ These include “limited training in palliative and end of life care, poor integration with wider health and social care systems, and restricted access to patient records, essential medicines and clinical advice.”^28^ Our findings support calls for the need to prioritise palliative and end-of-life care within ambulance services, including routine access to specialist advice and alternative care pathways where appropriate.

People in their last year of life account for fewer than 1 in 10 ED visits. While this represents an important patient group, their relatively small contribution to overall ED activity highlights the challenge of prioritising palliative and end-of-life care within a setting facing multiple competing demands. These challenges are compounded by issues such as a lack of training in palliative care among emergency care staff and lack of access to specialist advice, and health system restrictions including bed availability and availability of community services to support discharge planning.^29^

Our findings add to a growing body of evidence demonstrating that major health system shocks can alter patterns of end-of-life care and place additional pressure on services caring for people approaching death.^24,30^ Previous studies have documented increased palliative care needs alongside marked changes in care delivery during the Covid-19 pandemic.^31,32^ End-of-life care should be recognised as a core component of health system resilience planning.

## Conclusion

Approximately one-third of unplanned bed days are for people in their last year of life indicating that care of the dying is a core activity for hospital. Almost one in five ambulance conveyances to the ED are for people in their last year of life, highlighting a need to prioritise palliative and end of life care in ambulance services. Together, these findings show that people in their last year of life account for a substantial proportion of demand for acute hospital and ambulance services. The changes observed during the Covid-19 pandemic underscores the need to consider end-of-life care needs when planning and delivering acute hospital services during periods of exceptional pressure. Improving the quality and coordination of end-of-life care across hospital and community care services should be a key consideration in efforts to shift care from hospital to community and meet the needs of an ageing population while managing pressures on the health system.

## Contributors

JD, KS, AB and JV conceived and designed the study. TB and JL curated the data. JD carried out the analysis and drafted the initial manuscript. KS AB and JV provided input on statistical analysis. KS AB JV and RC provided input on interpretation and implications. All authors critically reviewed and edited the submitted manuscript, and all authors approved the final submitted manuscript. JD and KS are the guarantors.

The corresponding author attests that all listed authors meet authorship criteria and that no others meeting the criteria have been omitted.

## Patient and public involvement

Patients and/or the public were involved in the design, reporting, and dissemination plans of this research. Refer to the Methods section for further details.

## Patient consent for publication

Not applicable.

## Ethics approval

The North East-Newcastle and North Tyneside 2 research ethics committee provided ethical approval for the CVD-COVID-UK/COVID-IMPACT research programme (REC No 20/NE/0161) to access, within secure trusted research environments, unconsented, whole population, anonymised data from electronic health records collected as part of patients’ routine healthcare.

## Data Availability

All data produced in the present study are available upon application to the CVD-COVID-UK/COVID-IMPACT research programme.

https://github.com/BHFDSC/CCU110_01

## Acknowledgements

This work was funded by Marie Curie (MCCRP-24-06). KES is the Laing Galazka Chair at King’s College London, funded through an endowment from Cicely Saunders International and the Kirby Laing Foundation. The study was carried out with the support of the BHF Data Science Centre led by Health Data Research UK (BHF Grant no. SP/19/3/34678). This study made use of anonymised data held in NHS England’s Secure Data Environment service for England and made available via the BHF Data Science Centre’s CVD-COVID-UK/COVID-IMPACT consortium. This work used data provided by patients and collected by the NHS as part of their care and support. We would also like to acknowledge all data providers who make health relevant data available for research.

The British Heart Foundation Data Science Centre (grant No SP/19/3/34678, awarded to Health Data Research (HDR) UK) funded co-development (with NHS England) of the Secure Data Environment service for England, provision of linked datasets, data access, user software licences, computational usage, and data management and wrangling support, to coordinate national COVID-19 priority research. Consortium partner organisations funded data access, user software licences and computational usage, and the time of contributing data analysts, biostatisticians, epidemiologists, and clinicians.

## Data sharing statement

The data used in this study are available in NHS England’s Secure Data Environment (SDE) service for England, but as restrictions apply they are not publicly available (https://digital.nhs.uk/services/secure-data-environment-service). The CVD-COVID-UK/COVID-IMPACT research programme, led by the BHF Data Science Centre (https://bhfdatasciencecentre.org/), received approval to access data in NHS England’s SDE service for England from the Advisory Group for Data (AGD) (https://digital.nhs.uk/about-nhs-digital/corporate-information-and-documents/advisory-group-for-data) – formerly the Independent Group Advising on the Release of Data (IGARD) – via an application made in the Data Access Request Service (DARS) Online system (ref. DARS-NIC-381078-Y9C5K) (https://digital.nhs.uk/services/data-access-request-service-dars/dars-products-and-services). The CVD-COVID-UK/COVID-IMPACT Approvals & Oversight Board (https://bhfdatasciencecentre.org/areas/cvd-covid-uk-covid-impact/) subsequently granted approval to this project (CCU110) to access the data within NHS England’s SDE service for England. The anonymised data used in this study were made available to accredited researchers only. Those wishing to gain access to the data should follow the application process of the relevant national data custodian.

## Code sharing

The analysis using data in NHS Digital’s TRE for England was performed according to a pre specified analysis plan published on GitHub, along with the analysis code https://github.com/BHFDSC/ CCU110_01).

## Transparency statement

The lead author (the manuscript’s guarantor) affirms that the manuscript is an honest, accurate, and transparent account of the study being reported; no important aspects of the study have been omitted.

## Copyright/licence for publication

The Corresponding Author has the right to grant on behalf of all authors and does grant on behalf of all authors, a worldwide licence to the Publishers and its licensees in perpetuity, in all forms, formats and media (whether known now or created in the future), to i) publish, reproduce, distribute, display and store the Contribution, ii) translate the Contribution into other languages, create adaptations, reprints, include within collections and create summaries, extracts and/or, abstracts of the Contribution, iii) create any other derivative work(s) based on the Contribution, iv) to exploit all subsidiary rights in the Contribution, v) the inclusion of electronic links from the Contribution to third party material where-ever it may be located; and, vi) licence any third party to do any or all of the above.

## References

1. Morley C, Unwin M, Peterson GM, Stankovich J, Kinsman L. Emergency department crowding: A systematic review of causes, consequences and solutions. PLoS One. 2018;13(8):e0203316.

2. Ruxin T, Feldmeier M, Addo N, Hsia RY. Trends by Acuity for Emergency Department Visits and Hospital Admissions in California, 2012 to 2022. JAMA Network Open. 2023;6(12):e2348053–e2348053.

3. Alkhatib A, Aylin P, Klaber R, Woodcock T. Shifting patterns in emergency department attendance: a time series analysis. Emerg Med J. 2026;43(4):223.

4. Mason B, Kerssens JJ, Stoddart A, et al. Unscheduled and out-of-hours care for people in their last year of life: a retrospective cohort analysis of national datasets. BMJ Open. 2020;10(11):e041888.

5. Davies JM, Leniz J, Chua KC, et al. Association between ethnicity and emergency department visits in the last three months of life in England: a retrospective population-based study using electronic health records. BMJ Public Health. 2024;2(2):e001121.

6. Ma JE, Olsen MK, McDermott CL, et al. Factors Associated With Hospital Admission in the Last Month: A Retrospective Single Center Analysis. J Pain Symptom Manage. 2024;67(6):535–543.

7. Assareh H, Stubbs JM, Trinh LTT, Muruganantham P, Jalaludin B, Achat HM. Variation in Hospital Use at the End of Life Among New South Wales Residents Who Died in Hospital or Soon After Discharge. J Aging Health. 2020;32(7-8):708–723.

8. Cardona-Morrell M, Kim JCH, Brabrand M, Gallego-Luxan B, Hillman K. What is inappropriate hospital use for elderly people near the end of life? A systematic review. Eur J Intern Med. 2017;42:39–50.

9. Chambers D, Cantrell A, Preston L, et al. Reducing unplanned hospital admissions from care homes: a systematic review. Health Soc Care Deliv Res. 2023;11(18):1– 130.

10. Chambers D, Cantrell A, Booth A. Implementation of interventions to reduce preventable hospital admissions for cardiovascular or respiratory conditions: an evidence map and realist synthesis. Southampton (UK): NIHR Journals Library; 2020 Jan. (Health Services and Delivery Research, No. 8.2.) Scientific summary. Available from: https://www.ncbi.nlm.nih.gov/books/NBK552337/.

11. Clark D, Armstrong M, Allan A, Graham F, Carnon A, Isles C. Imminence of death among hospital inpatients: Prevalent cohort study. Palliative medicine. 2014;28(6):474–479.

12. Pring A, Westwood J, Bowtell N, Verne J. 51 Emergency hospital admissions near the end of life. BMJ Supportive & Palliative Care. 2024;14(Suppl 1):A20–A21.

13. Mitchell S, Barclay S, Evans C, Sleeman K. Palliative and end-of-life care in primary care during the COVID-19 pandemic and beyond. Br J Gen Pract. 2022;72(714):6–7.

14. Sleeman KE LJ, Davies JM, Bone AE, Pask S, Chambers RL, Kumar R, Fraser LK, Hocaoglu MB, Oluyase AO, Barclay S, Murtagh FEM & Higginson IJ Better End of Life 2022. Fairer Care at Home. The Covid-19 pandemic: a stress test for palliative and end of life care. Research report. London (UK): Marie Curie. 14 July 2022. accesed (07/12/22): https://www.mariecurie.org.uk/globalassets/media/documents/research/publications/beol-2022/h903a-beol-england.pdf.2022.

15. Cummins L, Julian S, Georghiou T, Kumar G, Scobie S. 2025. Public expenditure in the last year of life. The Nuffield Trust. available: https://www.nuffieldtrust.org.uk/sites/default/files/2025-02/Public_expenditure_in_the_last%20year_of_life_finalreport.pdf accessed: 18.09.26.

16. Wood A, Denholm R, Hollings S, et al. Linked electronic health records for research on a nationwide cohort of more than 54 million people in England: data resource. BMJ. 2021;373:n826.

17. BHF Health Data Science Curated Asset for Demographics. available: https://bhfdsc.github.io/documentation/curated_assets/kpcs accessed: 24.09.26.

18. HES APC CIPS curated asset. available: https://webarchive.nationalarchives.gov.uk/ukgwa/20180307232845tf_/http://content.digital.nhs.uk/media/11859/Provider-Spells-Methodology/pdf/Spells_Methodology.pdf accessed: 24.09.26.

19. NHS Data Dictionary ’Admission Method’ available: https://www.datadictionary.nhs.uk/attributes/admission_method.html accessed: 24.09.26.

20. Office for National Statistics. June 2021. Coronavirus (COVID-19) Infection Survey technical article: waves and lags of COVID-19 in England. Available here: https://www.ons.gov.uk/peoplepopulationandcommunity/healthandsocialcare/conditionsanddiseases/articles/coronaviruscovid19infectionsurveytechnicalarticle/wavesandlagsofcovid19inenglandjune2021 accessed: 08.04.2025.

21. McLennan D, Noble S, Noble M, Plunkett E, Wright G, Gutacker N. 2019. The English Indices of Deprivation 2019, Technical report. *Ministry of Housing*, Communities and Local Government.

22. ICB 2022 boundaries available: https://geoportal.statistics.gov.uk/datasets/ons::lsoa-2011-to-sicbl-to-integrated-care-boards-to-lad-july-2022-lookup-in-en/explore accessed: 24.09.26.

23. Langan SM, Benchimol EI, Guttmann A, et al. Setting the RECORD straight: developing a guideline for the REporting of studies Conducted using Observational Routinely collected Data. Clin Epidemiol. 2013;5:29–31.

24. Leniz J, Davies JM, Bone AE, et al. Deaths at home, area-based deprivation and the effect of the Covid-19 pandemic: An analysis of mortality data across four nations. Palliative medicine. 2023;37(7):1034–1039.

25. Marmot M, Allen, J., Boyce, T., Goldblatt, P., Morrison, J. Health equity in England: The Marmot Review 10 years on. London: Institute of Health Equity; 2020.

26. Johansson T, Sleeman KE, Finucance A, et al. Unmet palliative care needs in England and Wales: population-based estimates and future projections (2025-2050). medRxiv. 2026:2026.2002.2016.26345914.

27. National Audit of Care at the End of Life (NACEL) 2025. available: https://www.nacel.nhs.uk/patients-and-carers-tool-data accessed: 18.09.26.

28. Campling, N., Anstee, S., Latter, S., Richardson, A., Turnbull, J., Scott-Green, J. and Voss, S. (2025) Enhancing Paramedic Care at End of Life, Policy Brief, University of Southampton. available: 10.5258/SOTON/P1246 accessed: 18.09.26.

29. Sausman J, Arif A, Young A, et al. Healthcare professionals’ perspectives of the management of people with palliative care needs in the emergency department of a UK hospital. BMC Palliat Care. 2023;22:129.

30. Bajwah S, Koffman J, Hussain J, et al. Specialist palliative care services response to ethnic minority groups with COVID-19: equal but inequitable—an observational study. BMJ Supportive & Palliative Care. 2024;14(e1):e1478.

31. Connolly M, Bell M, Lawler F, Timmins F, Ryder M. Hospital-Based Palliative and End-of-Life Care in the COVID-19 Pandemic: A Scoping Review. Am J Hosp Palliat Care. 2022;39(9):1105–1120.

32. Jackson M. British medical bulletin article: resourcing of palliative and end of life care in the UK during the Covid-19 pandemic. Br Med Bull. 2022;142(1):44–51.

